# Association of Mass Distribution of Rapid Antigen Tests and SARS-CoV-2 Prevalence: Results from NIH-CDC funded Say Yes! Covid Test program in Michigan

**DOI:** 10.1101/2022.03.26.22272727

**Authors:** Apurv Soni, Carly Herbert, Jonggyu Baek, Qiming Shi, Juan Marquez, Emma Harman, Vik Kheterpal, Chris Nowak, Thejas Suvarna, Honghuang Lin, William Heetderks, Adrian Zai, Michael Cohen-Wolkowiez, Giselle Corbie-Smith, Warren Kibbe, Ben S. Gerber, Nathaniel Hafer, Bruce Barton, John Broach, David McManus

**Affiliations:** Program in Digital Medicine, Department of Medicine, University of Massachusetts Chan Medical School, Worcester, MA, USA; Division of Clinical Informatics, Department of Medicine, University of Massachusetts Chan Medical School, Worcester, MA, USA; Department of Population and Quantitative Health Sciences, University of Massachusetts Chan Medical School, Worcester, MA, USA; University of Massachusetts Center for Clinical and Translational Science, University of Massachusetts Chan Medical School, Worcester, MA, USA; Washtenaw County Health Department, Washtenaw, MI, USA; CareEvolution LLC, Ann Arbor, MI, USA; National Institute of Biomedical Imaging and Bioengineering, NIH, via contract with Kelly Services, Bethesda, MD, USA; Department of Pediatrics, Duke University School of Medicine, Durham, NC, USA; Center for Health Equity Research, Department of Social Medicine, Department of Medicine, University of North Carolina School of Medicine, Chapel Hill, NC, USA; Department of Biostatistics and Bioinformatics, Duke University School of Medicine, Durham, NC, USA; Department of Emergency Medicine, University of Massachusetts Chan Medical School, Worcester, MA, USA; Division of Cardiology, Department of Medicine, University of Massachusetts Chan Medical School, Worcester, MA, USA

## Abstract

**Importance:** Wide-spread distribution of diagnostics is an integral part of the United States’ COVID-19 strategy; however, few studies have assessed the effectiveness of this intervention at reducing transmission of community COVID-19.

**Objective:** To assess the impact of the Say Yes! Covid Test (SYCT!) Michigan program, a population-based program that distributed 20,000 free rapid antigen tests within Ann Arbor and Ypsilanti, Michigan in June-August 2021, on community prevalence of SARS-CoV-2.

**Design:** This ecological study analyzed cases of SARS-CoV-2 from March to October 2021 reported to the Washtenaw County Health Department.

**Setting:** Washtenaw County, Michigan

**Participants:** All residents of Washtenaw County

**Interventions:** Community-wide distribution of 500,000 rapid antigen tests for SARS-CoV-2 to residents of Ann Arbor and Ypsilanti, Michigan. Each household was limited to one test kit containing 25 rapid antigen tests.

**Main Outcome and Measures:** Community prevalence of SARS-CoV-2, as measured through 7-day average cases, in Ann Arbor and Ypsilanti was compared to the rest of Washtenaw County. A generalized additive model was fitted with non-parametric trends for control and relative differences of trends in the pre-intervention, intervention, and post-intervention periods to compare intervention municipalities of Ann Arbor and Ypsilanti to the rest of Washtenaw County. Model results were used to calculate average cases prevented in the post-intervention period.

**Results:** In the post-intervention period, there were significantly lower standardized average cases in the intervention communities of Ann Arbor/Ypsilanti compared to the rest of Washtenaw County (p<0.001). The estimated standardized relative difference between Ann Arbor/Ypsilanti and the rest of Washtenaw County was -0.016 cases per day (95% CI: -0.020 to -0.013), implying that the intervention prevented 40 average cases per day two months into the post-intervention period if trends were consistent.

**Conclusions and Relevance:** Mass distribution of rapid antigen tests may be a useful mitigation strategy to combat community transmission of SARS-CoV-2, especially given the recent relaxation of social distancing and masking requirements.

## Introduction

Large scale public health efforts, including non-pharmaceutical mitigation strategies of testing, social distancing, and masking, remain the focal point of the national strategy to address the SARS-CoV-2 pandemic in the third year of this crisis.^1^ Government-supported programs to distribute free rapid antigen tests, including the federal program to distribute up to 500 million tests, are an important step towards this goal.^2^ While previous efforts of mass distribution of rapid antigen tests have been successful in demonstrating the feasibility of such programs, none have demonstrated the program’s effectiveness at reducing transmission of COVID-19 in the community.^3,4^ This manuscript describes an ecological assessment of the Say Yes! Covid Test (SYCT!) Michigan program, a population-based program that distributed 500,000 free rapid antigen tests within Ann Arbor and Ypsilanti, Michigan in June-August 2021, on SARS-CoV-2 community prevalence.

## Methods

The Say Yes! Covid Test Michigan program distributed 500,000 free rapid antigen tests between June 4 and August 11, 2021 to the residents of Ann Arbor and Ypsilanti, two municipalities in Washtenaw County, Michigan (Figure 1). Combined, Ann Arbor and Ypsilanti account for approximately 40% of total residents in Washtenaw County (Supplemental Table 1). The program was administered by the National Institutes of Health and Centers for Disease Control and Prevention, in partnership with the Washtenaw County Health Department. The tests were distributed via online ordering and direct shipment to resident’s homes and by local distribution through pick-up sites at schools, churches, and community events. Each household was limited to one test kit containing 25 rapid antigen tests, and 20,000 kits were distributed in total. More information about the design and implementation of the SYCT! program can be found elsewhere.^3,4^ We evaluated 7-day moving average cases of SARS-CoV-2 in Ann Arbor and Ypsilanti vs. the rest of Washtenaw County during the intervention (June 4-August 11, 2021) and post-intervention (August 11-October 15, 2021) periods. Municipality-level SARS-CoV-2 case numbers in Washtenaw County were obtained through the Washtenaw County Health Department. Case-rates in the pre-intervention period (March 1-June 3, 2021) were standardized to adjust for pre-intervention differences of trends. To test the trends in the post-intervention period, a generalized additive model (GAM) with spline terms of pre-intervention, intervention, and post-intervention was fitted. Specifically, the fitted GAM for Ann Arbor/Ypsilanti compared to the rest of Washtenaw County was:

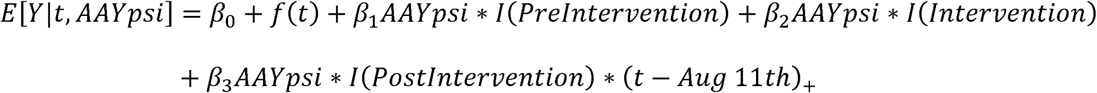

where *l(Prelntervention), l(lntervention)*, and *l(Postlntervention)* are indicators for time in the pre-intervention, intervention, post-intervention period, respectively, *AAYpsi* is an indicator for Ann Arbor and Ypsilanti, and *(t - Aug* 11*th)*_+_ is a linear spline slope term in the post intervention period. A model was also generated to compare the separate trends in the post-intervention period in Ypsilanti and Ann Arbor. For this model, functional terms of patterns were included: a quadratic trend difference for Ann Arbor and a linear trend difference for Ypsilanti. In the fitted GAM models, degree of smoothness was chosen by the automatic selection using generalized cross-validation in gam() function in mgcv R package.^5^ Model estimates were used to calculate average cases prevented in the post-intervention period. All analyses were conducted in R 4.1.1.^6^ The SYCT! Michigan intervention received non-research determination by the University of Massachusetts Chan Medical School Institutional Review Board.

**Figure 1:**
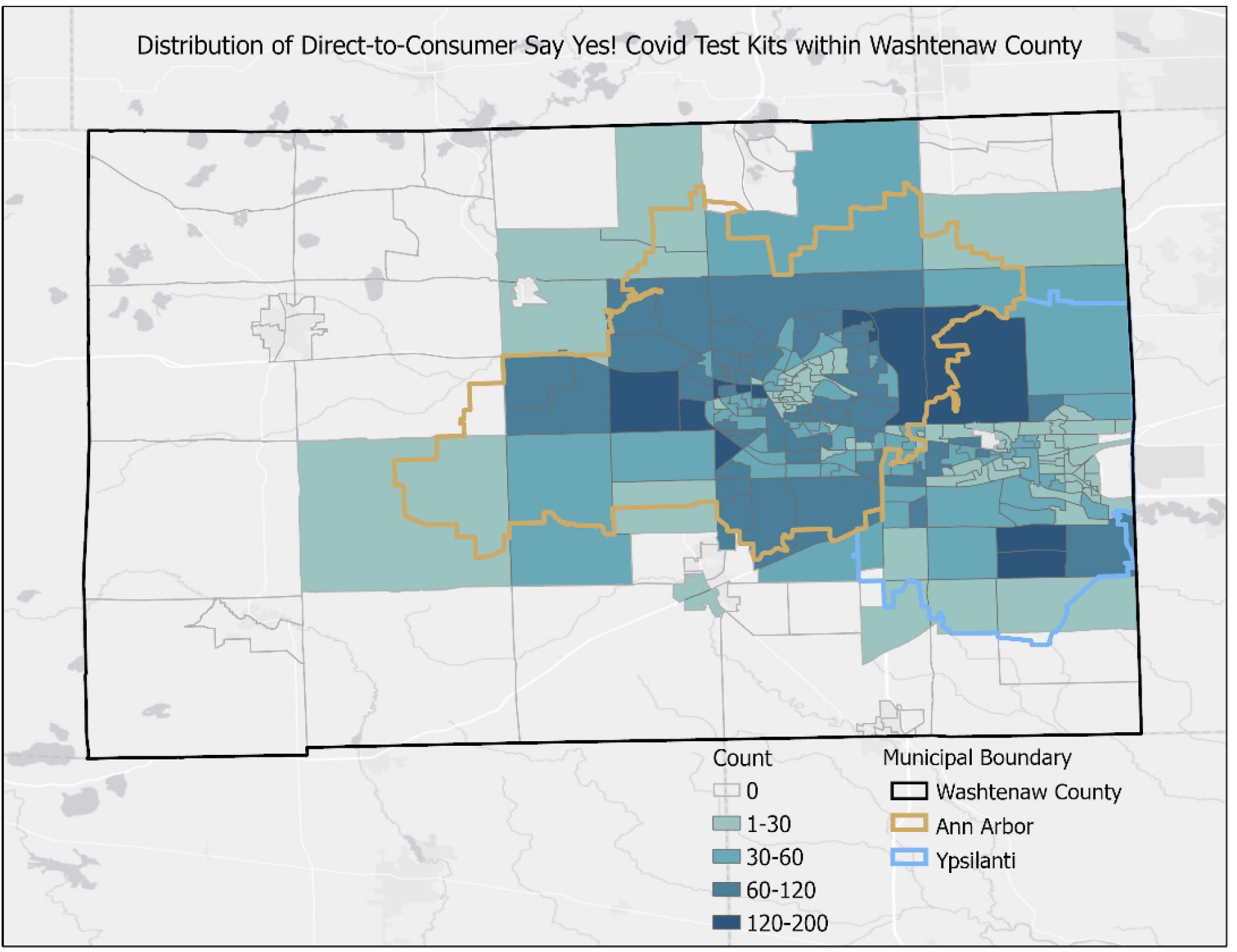
Distribution of Direct-to-Consumer Say Yes Covid Test Kits in Washtenaw County.

## Results

Unadjusted cases in Ann Arbor and Ypsilanti compared to the rest of Washtenaw County are shown in Supplemental Figure 1. After standardizing for variance in the pre-intervention period, we found no significant difference in SARS-CoV-2 cases in the intervention period between Ann Arbor and Ypsilanti compared to the rest of Washtenaw County. However, in the post-intervention period, as cases increased across all communities, there were significantly lower standardized average cases in Ann Arbor and Ypsilanti compared to the rest of Washtenaw County (p<0.001) (Figure 2a; Supplemental Figure 2). The estimated standardized relative difference between Ann Arbor/Ypsilanti and the rest of Washtenaw County was -0.016 cases per day (95% CI: -0.020 to -0.013). We estimate that the intervention prevented 40 average cases per day two months into the post-intervention period if trends of the average cases between Ann Arbor/Ypsilanti and the rest of Washtenaw were consistent (Supplementary Table 1, Supplementary Table 2). Separate evaluation of Ann Arbor and Ypsilanti average cases revealed a similar pattern, though the intervention had a delayed effect in Ann Arbor compared to Ypsilanti (Figure 2b), attributable to a spike in cases in Ann Arbor during late August/early September.

**Figure 2:**
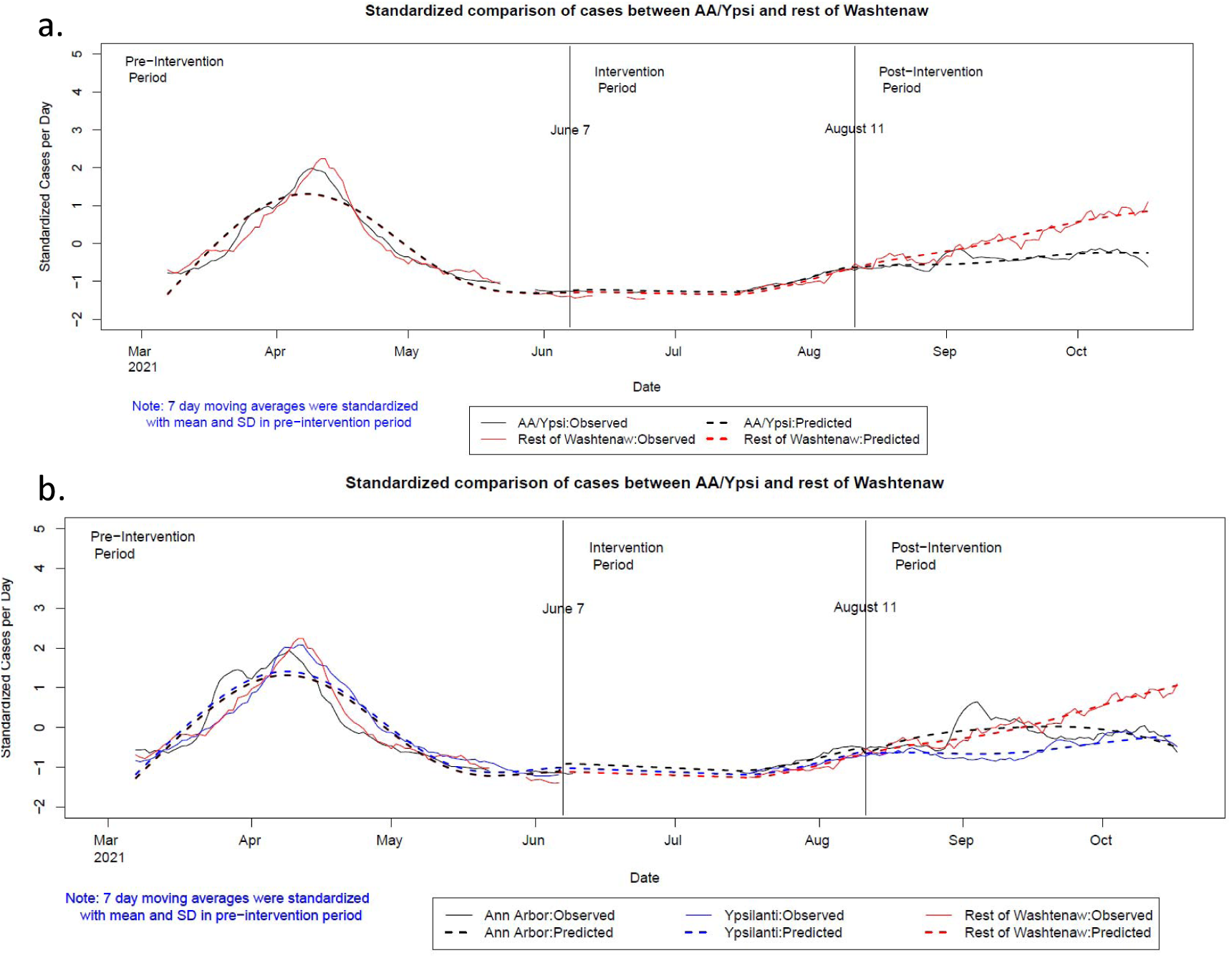
Standardized Comparison of 7-day Moving Average Cases of SARS-CoV-2 infections and predicted cases in Ann Arbor and Ypsilanti compared to the rest of Washtenaw County, March-September 2021. Footnote: Predicted cases according to fitted generalized additive model. The smoothness of the fitted generalized additive model was chosen automatically based on generalized cross-validation.

## Discussion

In this evaluation of the large-scale distribution of rapid antigen tests in two Michigan municipalities, we observed a significantly lower average case-rate in intervention municipalities two months after test distribution. This finding was observed in the context of the Delta-variant related SARS-CoV-2 surge in the state of Michigan. With the relaxation of indoor mask guidelines by the Centers for Disease Control and Prevention, timely testing will be a centerpiece of non-pharmaceutical mitigation strategy.^7^ In that context, these findings suggest that mass distribution of rapid antigen tests is a viable strategy for reducing transmission of SARS-CoV-2, especially during a surge.

These findings must be interpreted in the context of the ecological design, which limits our ability to draw causal inferences. Consistent effects were observed in both municipalities that received the tests, despite marked differences between municipalities in vaccination rates before and after the intervention period, suggesting the observed reduction in cases is not entirely attributable to differences in vaccination rates (Supplemental Table 1). It is important to note that the Washtenaw County Health Department COVID-19 data is predominantly comprised of molecular (PCR) test results, and rapid antigen tests are often unreported to health departments. Therefore, observed differences may be attributable to a reporting bias such that residents in intervention towns, where rapid antigen tests were more widely available due to the SYCT! Michigan intervention, performed molecular tests less frequently. Unfortunately, data for total number of tests performed is not available from Washtenaw County Health Department at the sub-county level, precluding our ability to test this assumption. However, we present robust findings using rigorous non-parametric methods from the best data available. Finally, this program distributed 25 tests to each household, which is 6-fold higher than the number of tests being distributed by the US government throughout the country. Therefore, caution must be applied for generalizing findings from this study to possible effectiveness of the federal program.

### Public Health Implications and Conclusions

Rapid antigen tests offer unique advantages over more sensitive real-time reverse transcriptase polymerase chain reaction (RT-PCR) tests as a public health strategy including relatively low cost, ease of use, and availability of results in 15 minutes.^8^ These benefits need to be weighed against the limitations of rapid antigen tests, including lower sensitivity among asymptomatic individuals and those with low viral load.^8,9^ Our finding indicating that distribution of tests several months prior to a surge in SARS-CoV-2 cases was still associated with a reduction in community transmission suggests that recipients of this intervention were responsible owners of a personal supply of rapid antigen tests. These findings, taken together with recent reports that rapid antigen tests’ performance remain consistent across variants, underscore the continued utility of rapid antigen tests.^9^ With emerging reports of the BA.2 variant causing a surge in cases across the globe and in the United States, these findings hold particular importance for the public health of the country.

## Data Availability

All data produced in the present study are available upon reasonable request to the authors.

## Acknowledgment

We are deeply grateful first to the local community of Washtenaw County Michigan, including the local health officials and second to our collaborators from the National Institute of Health (NIBIB and NHLBI) who provided scientific input into the design of this study and interpretation of our results, but could not formally join as co-authors due to institutional policies. We received meaningful contributions from Drs. Bruce Tromberg, Jill Heemskerk, Rachael Fleurence, Andrew Weitz, Krishna Juluru, Felicia Qashu, Dennis Buxton, Jue Chen and Erin Itturriaga.

## Figures and Tables

**Table 1:** Population, SARS-CoV-2 Cases, and Vaccination Rate in Ann Arbor and Ypsilanti, June-October 2021.

| Region | Population (2019): | Mean SARS-CoV-2 Cases in the Pre-Period<br>n (SD) | Vaccination Rate<sup>b</sup>: Start of Intervention (June 7<sup>th</sup>) | Vaccination Rate: End of Intervention (August 11<sup>th</sup>) | Vaccination Rate: Post-Intervention (October 15<sup>th</sup>) |
| --- | --- | --- | --- | --- | --- |
| **Ann Arbor** | 120,735 | 25.2 (20.2) | 72.4 | 75.3 | 78.3 |
| **Ypsilanti** | 20,828 | 27.6 (21.0) | 54.3 | 59.1 | 62.8 |
| **Washtenaw County<sup>a</sup>** | 226,038 | 23.1 (39.8) | 62.0 | 65.2 | 67.7 |

<sup>a</sup>Excluding Ann Arbor and Ypsilanti
<sup>b</sup>Percent of individuals who have received 1+ doses of a SARS-CoV-2 vaccine

