## Additional Methods for "Association of Mass Distribution of Rapid Antigen Tests and SARS-CoV-2 Prevalence: Results from NIH-CDC funded Say Yes! Covid Test program in Michigan"

**Data Sources:**

*Data Source for Figure 1*: All direct-to-consumer (DTC) Say Yes! Covid Test kit orders were processed through an online platform developed by CareEvolution. Number of DTC orders were recorded at the zip-code level. All orders were deidentified using anonymous participant identifiers.

*Data Source for Ecological Analysis:* Municipality-level SARS-CoV-2 case numbers in Washtenaw County were obtained through the Washtenaw County Health Department.

**Statistical Analysis:**

To smooth out daily fluctuation of cases, 7-day moving averages of cases were calculated and standardized using mean and standard deviation in the pre-intervention period to adjust for differences of trends observed in the pre-intervention period. To test the different trends in the post-intervention period and to check the adjustment for the differences in the pre-intervention was successful by standardization, a generalized additive model (GAM) with spline terms of pre-intervention, intervention, and post-intervention was fitted. Specifically, the fitted GAM for Ann Arbor/Ypsilanti combined compared to the rest of Washtenaw County was:

$$E\left[ Y|t, AAYpsi \right]=\beta_{0}+f\left( t \right)+\beta_{1}AAYpsi*I\left( PreIntervention \right)+\beta_{2}AAYpsi*I\left( Intervention \right)+\beta_{3}AAYpsi*I\left( PostIntervention \right)*\left( t-Aug 11th \right)_{+}$$

where $I\left( PreIntervention \right)$, $I\left( Intervention \right)$, and $I\left( PostIntervention \right)$, are an indicator for time in the pre-intervention, intervention, post-intervention period, respectively; $AAYpsi$ is an indicator for Ann Arbor and Ypsilanti; $\left( t-Aug 11th \right)_{+}$ is a linear spline slope term in the post intervention period to model a slope difference between Ann Arbor/Ypsilanti and the rest of Washtenaw in the post-intervention period. This spline term $\left( t-Aug 11th \right)_{+}=0$ before or on Aug 11^th^, 1 on Aug 12^th^, and so on. The baseline trends $\beta_{0}+f\left( t \right)$ non-parametrically estimates the average cases in the rest of Washtenaw over time; regression parameters $\beta_{1}$, and $\beta_{2}$ are the difference of the average cases between Ann Arbor/Ypsilanti and the rest of Washtenaw for the pre-intervention and intervention periods, respectively; and $\beta_{3}$ is a slope difference (i.e., the average difference per day) between Ann Arbor/Ypsilanti and the rest of Washtenaw in the post-intervention period. In the fitted GAM models, degree of smoothness was chosen by the automatic selection using generalized cross-validation in gam() function in mgcv R package. A sensitivity analysis with a fixed degree of smoothness specifying knot k=15 was performed to check the consistency of the estimate for $\beta_{3}$ (Supplemental Figure 2).

Number of cases prevented in the post-intervention period in Ann Arbor and Ypsilanti combined was calculated by the following equation:

$$Prevented cases\left( combined AAYpsi \right) at time t=\beta_{3}*t*{SD}_{AAYpsi}$$

Where *t* is number of days in the post-intervention period and *SD* is standard deviation in the pre-period.

A separate model was also generated to compare the trends in the post-intervention period in Ypsilanti and Ann Arbor, separately. For this model analyzing the difference between municipalities, functional terms of patterns were included: a quadratic trend difference between Ann Arbor and the rest of Washtenaw and a linear trend difference between Ypsilanti and the rest of Washtenaw. This GAM model was specified as follows:

$E\left[ Y|t, AA,Ypsi \right]=\beta_{0}+f\left( t \right)+\beta_{1}AA*I\left( PreIntervention \right)+\beta_{2}Ypsi*I\left( PreIntervention \right)+\beta_{3}AA*I\left( Intervention \right){+ \beta}_{4}Ypsi*I\left( Intervention \right)+ \beta_{5}AA*I\left( PostIntervention \right)\left( t-Aug 11th \right)_{+}+ \beta_{6}AA*I\left( PostIntervention \right)\left( t-Aug 11th \right)_{+}^{2}+ \beta_{7}Ypsi*I\left( PostIntervention \right)\left( t-Aug 11th \right)_{+}$

Number of cases prevented in the post-intervention period was calculated by the following equations:

$$Prevented cases\left( AA \right) at time t={(\beta}_{5}*{SD}_{AA}) t+{(\beta}_{6}*{SD}_{AA})t^{2}$$

$$Prevented cases\left( Ypsi \right) at time t={(\beta}_{7}*{SD}_{Ypsi}) t$$

Where *t* is number of days in the post-intervention period and *SD* is standard deviation in the pre-period.

Methods Citations:

1.	Ciccone EJ, Conserve DF, Dave G, et al. At-home testing to mitigate community transmission of SARS-CoV-2: protocol for a public health intervention with a nested prospective cohort study. *BMC Public Health*. 2021;21(1):1-15. doi:10.1186/S12889-021-12007-W/TABLES/3

2.	CDC–NIH Initiative Provides Free COVID-19 Rapid Home Tests In North Carolina, Tennessee | Health Affairs. Accessed February 28, 2022. https://www.healthaffairs.org/do/10.1377/forefront.20211025.437195/full/

3.	Wood S. Mixed GAM Computation Vehicle with Automatic Smoothness Estimation. Published online February 24, 2022.
