## Supplemental Figures and Tables for "Association of Mass Distribution of Rapid Antigen Tests and SARS-CoV-2 Prevalence: Results from NIH-CDC funded Say Yes! Covid Test program in Michigan"

**Supplementary Table 1. Relative average SARS-COV-2 case differences in Ann Arbor/Ypsilanti compared to the rest of Washtenaw County.**

|  | Time Period | Average difference in cases per day^a^ | 95% Confidence Interval | | P-value |
| --- | --- | --- | --- | --- | --- |
| Ann Arbor and Ypsilanti Combined | Overall difference in pre-intervention | 0.581 | -2.335 | 3.497 | 0.697 |
|  | Overall difference in intervention | 2.463 | -1.697 | 6.624 | 0.247 |
|  | Average difference per day in post-intervention | -0.654 | -0.739 | -0.569 | <0.001 |
| Ann Arbor | Overall difference in pre-intervention | 0.070 | -1.594 | 1.733 | 0.934 |
|  | Overall difference in intervention | 3.694 | 1.044 | 6.345 | 0.007 |
|  | Average difference per day (i.e., a linear spline slope term) in post-intervention | 0.478 | 0.310 | 0.647 | <0.001 |
|  | Average difference per day (i.e., a quadratic spline slope term) in post-intervention | -0.014 | -0.017 | -0.011 | <0.001 |
| Ypsilanti | Overall difference in pre-intervention | 2.007 | 0.309 | 3.705 | 0.021 |
|  | Overall difference in intervention | 1.440 | -1.529 | 4.408 | 0.342 |
|  | Average difference per day (i.e., a linear spline slope term) in post-intervention | -0.390 | -0.440 | -0.340 | <0.001 |

^a^Regression estimates from fitted generalized additive models (GAMs) were multiplied by standard deviation of cases in pre-intervention period.

**Supplementary Table 2. SARS-CoV-2 Cases Prevented per Day by Say Yes! Covid Test Michigan Intervention Two Months Post-Intervention**

| Region | Cases Prevented per Day  (at Day 60) |
| --- | --- |
| Ann Arbor | 22.1 |
| Ypsilanti | 23.4 |
| Ann Arbor/Ypsilanti combined | 39.2 |

**Supplemental Figure 1: Unadjusted Cases per Day of SARS-CoV-2 in Ann Arbor (AA) and Ypsilanti (Ypsi) compared to the rest of Washtenaw County**

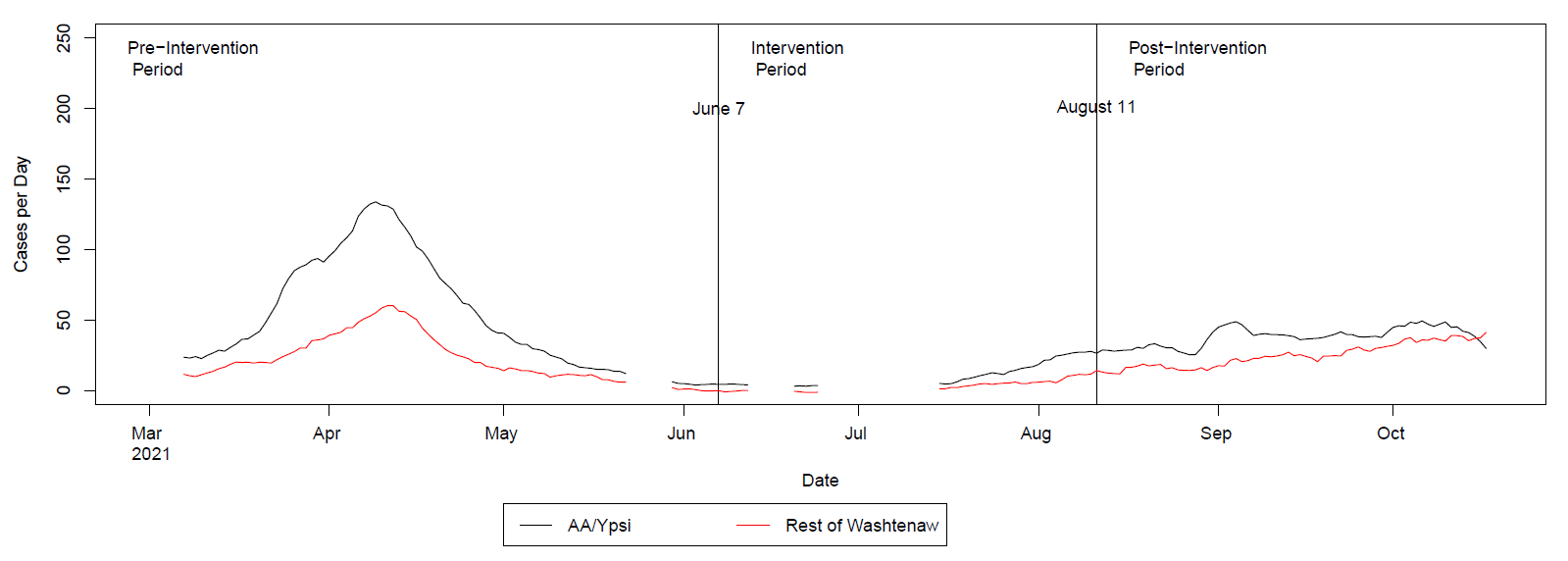

**Supplemental Figure 2: Standardized Comparison of 7-day Moving Average SARS-CoV-2 infections in Ann Arbor (AA) and Ypsilanti (Ypsi) compared to the rest of Washtenaw County, March-September 2021, after adjusting for over-smoothness in pre-intervention period**

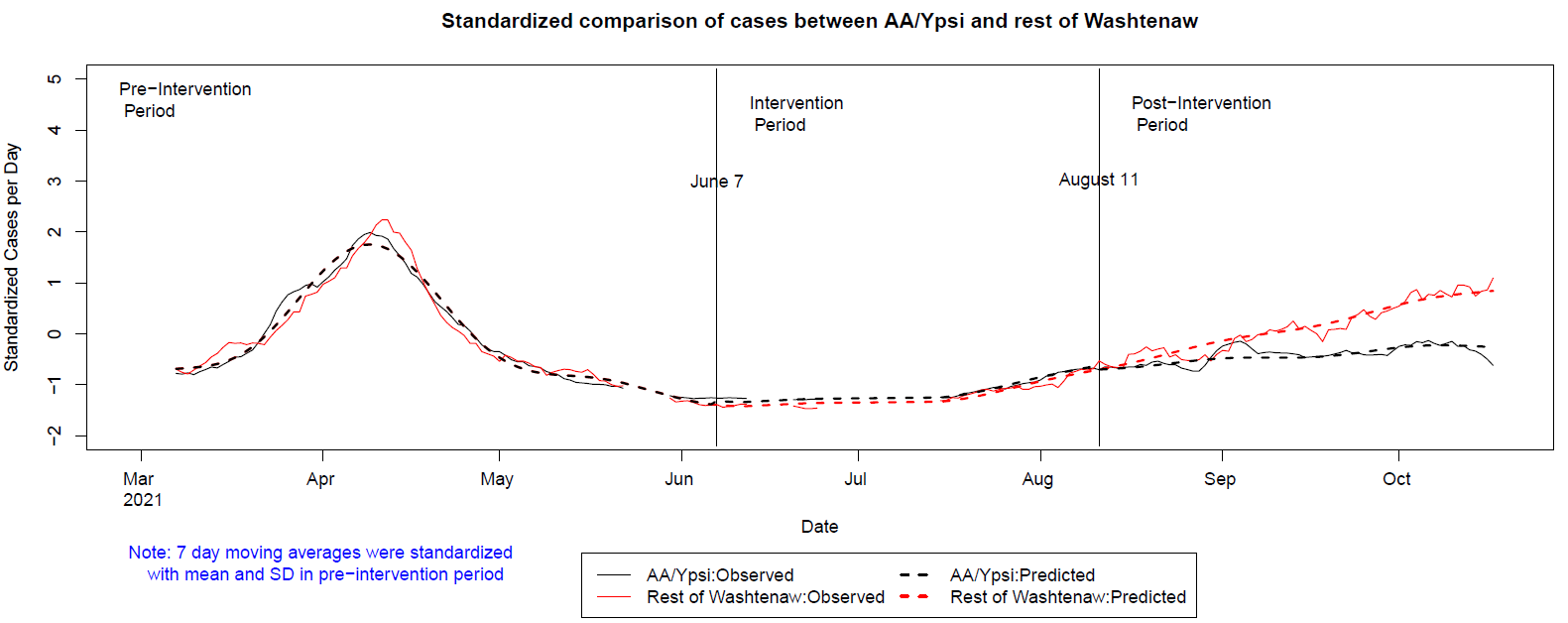

Footnote: The smoothness of the fitted GAM was chosen with specifying fixed degree of smoothness k=15 to adjust for over-smoothness of predicted trends in pre-intervention period.
